# The role of multi-generational household clusters in COVID-19 in England

**DOI:** 10.1101/2021.11.22.21266540

**Authors:** Simon Thelwall, Asad Zaidi, Olisaeloka Nsonwu, Wendy Rice, Dimple Chudasama, Theresa Lamagni, Gavin Dabrera

## Abstract

**Background:** Household transmission has been demonstrated to be an important factor in the population-level growth of COVID-19. UK Health Security Agency (UKHSA) maintains data on positive tests for COVID-19 and the residential addresses of cases. We sought to use this information to characterise clusters of COVID-19 in multi-generational households in England.

**Methods:** Using cross-sectional design, cases of COVID-19 were assigned to clusters if they occurred in the same residential property in a 14-day rolling window. Patient demographic data were supplemented with reference to the ONS index of multiple deprivation and population density. Multi-generational households were defined as a cluster with at least three people, with one case in a person who was 0-16 years old and one case in a person who was ≥ 60 years old, with at least 16 years between two members of each age group.

**Results:** A total of 3,647,063 COVID-19 cases were reported between 01 April 2020 and 20 May 2021. Of these, 1,980,527 (54.3 %) occurred in residential clusters. Multi-generational households formed 1.5 % of clusters, with these more likely to occur in areas of higher population density and higher relative deprivation. Multi-generational clusters were more common among households of non-White ethnicity and formed larger clusters than non-multi-generational clusters (median cluster size 6, IQR 4-11 vs 3, IQR 3-4, respectively).

**Conclusion:** Multi-generational clusters were not highly prevalent in England during the study period, however were more common in certain populations.

**BOX TEXT:** *What is already known on this subject:* Greater risk of infection with SARS-CoV-2 in England is associated with being of non-White ethnicity, residence in an area of greater deprivation and higher population density. What is less clear is the role of household composition in the risk of COVID-19 transmission. It has been hypothesised that multi-generational housing (in which at least three different generations are resident in the same property) accounted for a substantial proportion of COVID-19 cases. We sought to test this hypothesis.

*What this study adds:* This study provides descriptive evidence around the role of multi-generational households in the COVID-19 pandemic in England between April 2020 and March 2021. It does not support the hypothesis that this period (a period of low incidence in England), a substantial proportion of COVID-19 cases occurred in multi-generational households.

## INTRODUCTION

Household transmission is a high-risk route for infection and an important potential driver of COVID-19 incidence in the population. Analysis of the first 379 COVID-19 cases in England identified a secondary attack rate of 16% (95% CI 11-20) for confirmed secondary cases. [1,2]

The English experience of the COVID-19 pandemic in 2020 saw a summer nadir in case rates, raising questions about the settings in which transmission did occur. It was hypothesised that an important contributor to total case numbers was cases occurring in multi-generational households, and that multi-generational household clusters were more likely to be formed of individuals of non-white ethnicities.

There is no single definition of multi-generational households consistently used in research and statistics in England. One definition used three generations of adults in a single household and estimated 1.8 million such multi-generational households in the UK in 2013/14, a number that has increased since 2009/10. [4] Data from the Office for National Statistics’ (ONS) Annual Population Survey (APS) shows that multi-generational households (defined as households containing at least one individual aged 0-19, 20-69 and ≥ 70 years old) are more prevalent among non-White ethnicity households than among White ethnicity households. [5]

UK Health Security Agency (UKHSA) collects and maintains data on positive tests for SARS-CoV-2 including addresses where cases reside, as a central component of communicable disease surveillance. We sought to use this information to characterise outbreaks of COVID-19 in households in England to examine the hypothesis that cases of COVID-19 occurring in multi-generational households constituted a substantial proportion of the overall number of cases during the Summer nadir in cases in 2020.

## METHODS

We used data on all SARS-CoV-2 cases that tested positive by polymerase chain reaction (PCR) or lateral flow test (LFT) in England. Cases were deduplicated to the earliest positive test per person, in line with government reporting of COVID-19 data. [6]

Cases were address matched using Ordnance Survey AddressBase databases. The process preferentially used the patient address provided with the test. Where this was absent, the address held within the individual’s NHS Summary Care Record was used. This is the address held within a national NHS database derived from GP and other NHS healthcare records. Address matching returned a unique property reference numbers (UPRN) and Basic Land Property Unit (BLPU) class to determine the property classification. Cases occurring in properties other than private dwellings, such as care homes, medical facilities, prisons or university campuses, were excluded.

Clusters were defined as two or more cases occurring at the same residential property (UPRN), within a rolling 14-day window, starting with the earliest specimen date of the first laboratory confirmed case. In the absence of a standard pre-existing definition, multi-generational clusters were defined as those with three or more cases, with one case in a person who was 0-16 years old and one case in a person who was ≥ 60 years old, with at least 16 years between two members of each age group. Other definitions, which lack the requirement for an age gap between age groups, may be limited by the fact that two members of a household could have very similar ages but cross an age boundary, thus not truly representing different generations. For example the ONS definition requires three people with somebody aged 0-19, somebody 20-69 and somebody ≥ 70 years old. This could then be three people 19, 20 and 70 years old, which would represent only two generations rather than three.

Local-area deprivation associated with cases’ residential address was measured using the Index of Multiple Deprivation (IMD) using data from the Office for National Statistics, a composite score of deprivation across seven domains. These were analysed at lower layer super output area (LSOA), a census geography with a population between 1,000 and 3,000 people and 400 to 1,200 households. Population density data at the LSOA level was obtained from ONS. [7]

Self-reported ethnicity from test request data was used where available. Where unavailable, data were supplemented using data from linked-individual level data in Hospital Episode Statistics (HES), a database of all secondary care admissions for England.

Chi squared tests for independence were performed for categorical variables. Median tests were used to test the null hypothesis that there was no difference in medians for age and cluster size. Records with missing data for a given variable were reported as unknown. All hypothesis tests were performed with permutation testing using the R package coin. [8] All analyses were performed in R version 4.0.2 [9].

## RESULTS

Between 01 April 2020 and 20 May 2021 a total of 3,647,063 COVID-19 cases were reported in England. Of these, 1,980,527 (54.3 %) formed a total of 719,165 residential clusters across all settings. 96.5 % (n=694,109) of clusters occurred in private residential dwellings and 1.5 % of such clusters occurred in multi-generational households.

The median age of COVID-19 cases was slightly lower in multi-generational clusters, compared to non-multi-generational clusters (36 and 37 years respectively) and clusters tended to be larger (median cluster size 6 and 2 respectively, Table 1).

**Table 1:** Distribution of demographics by multi-generational clusters, England April 2020 to May 2021

| Demographic |  | Non-multi-generational clusters | Non-multi-generational clusters with at least three cases per cluster | Multi-generational clusters<sup>a</sup> | p |
| --- | --- | --- | --- | --- | --- |
| Median age | Years (IQR) | 37 (23 - 52) | 33 (19 - 48) | 36 (17 - 56) | < 0.001 |
| Median cluster size | Cases (IQR) | 2 (2 - 3) | 3 (3 - 4) | 6 (4 - 11) | < 0.001 |
| Mixed ethnicity status | Single | 575,467 (84.2%) | 184,192 (78.3%) | 6,349 (61.0%) | < 0.001 |
|  | Mixed | 108,241 (15.8%) | 51,035 (21.7%) | 4,052 (39.0%) |  |
| Ethnicity in single ethnicity households | Asian Bangladeshi | 10,719 (1.9%) | 5,146 (2.8%) | 360 (5.7%) | < 0.001 |
|  | Asian Indian | 30,262 (5.3%) | 13,658 (7.4%) | 1,123 (17.7%) |  |
|  | Asian other | 12,416 (2.2%) | 5,072 (2.8%) | 231 (3.6%) |  |
|  | Asian Pakistani | 30,021 (5.2%) | 13,596 (7.4%) | 1,025 (16.1%) |  |
|  | Black African | 11,230 (2.0%) | 3,980 (2.2%) | 142 (2.2%) |  |
|  | Black Caribbean | 4,001 (0.7%) | 1,037 (0.6%) | 38 (0.6%) |  |
|  | Black other | 574 (0.1%) | 105 (0.1%) | 1 (0.0%) |  |
|  | Mixed ethnicity | 3,111 (0.5%) | 668 (0.4%) | 17 (0.3%) |  |
|  | Other | 6,022 (1.0%) | 2,165 (1.2%) | 91 (1.4%) |  |
|  | Unknown | 13,983 (2.4%) | 3,885 (2.1%) | 115 (1.8%) |  |
|  | White British | 416,922 (72.4%) | 124,001 (67.3%) | 2,823 (44.5%) |  |
|  | White other | 36,206 (6.3%) | 10,879 (5.9%) | 383 (6.0%) |  |
| IMD quintile<sup>a</sup> | 1 | 159,108 (23.3%) | 55,197 (23.5%) | 2,882 (27.7%) | < 0.001 |
|  | 2 | 151,795 (22.2%) | 54,298 (23.1%) | 2,766 (26.6%) |  |
|  | 3 | 134,143 (19.6%) | 45,762 (19.5%) | 2,014 (19.4%) |  |
|  | 4 | 125,817 (18.4%) | 41,855 (17.8%) | 1,521 (14.6%) |  |
|  | 5 | 112,620 (16.5%) | 38,056 (16.2%) | 1,216 (11.7%) |  |
|  | Missing | 225 (0.0%) | 59 (0.0%) | 2 (0.0%) |  |

| Demographic |  | Non-multi-generational clusters | Non-multi-generational clusters with at least three cases per cluster | Multi-generational clusters* | p |
| --- | --- | --- | --- | --- | --- |
| Population density quintile<sup>b</sup> | 1 | 163,105 (23.9%) | 64,229 (27.3%) | 4,330 (41.6%) | < 0.001 |
|  | 2 | 147,282 (21.5%) | 50,616 (21.5%) | 2,035 (19.6%) |  |
|  | 3 | 136,692 (20.0%) | 44,976 (19.1%) | 1,528 (14.7%) |  |
|  | 4 | 131,743 (19.3%) | 42,435 (18.0%) | 1,430 (13.7%) |  |
|  | 5 | 104,661 (15.3%) | 32,912 (14.0%) | 1,076 (10.3%) |  |
|  | Missing | 225 (0.0%) | 59 (0.0%) | 2 (0.0%) |  |
| Region | East Midlands | 56,947 (8.3%) | 17,631 (7.5%) | 612 (5.9%) | < 0.001 |
|  | East of England | 78,596 (11.5%) | 28,493 (12.1%) | 1,004 (9.7%) |  |
|  | London | 130,144 (19.0%) | 54,373 (23.1%) | 4,054 (39.0%) |  |
|  | North East | 32,925 (4.8%) | 9,326 (4.0%) | 223 (2.1%) |  |
|  | North West | 105,501 (15.4%) | 32,171 (13.7%) | 925 (8.9%) |  |
|  | South East | 97,470 (14.3%) | 34,813 (14.8%) | 1,404 (13.5%) |  |
|  | South West | 36,993 (5.4%) | 11,907 (5.1%) | 337 (3.2%) |  |
|  | West Midlands | 78,366 (11.5%) | 26,718 (11.4%) | 1,159 (11.1%) |  |
|  | Yorkshire and Humber | 66,011 (9.7%) | 19,568 (8.3%) | 674 (6.5%) |  |
|  | Missing | 755 (0.1%) | 227 (0.1%) | 9 (0.1%) |  |

<sup>a</sup>1 = least deprived/, 5 = most deprived
<sup>b</sup>1 = highest density, 5 = lowest density
\*Hypothesis tests compare multigenerational clusters with non-multigenerational clusters with at least three cases per cluster

Multi-generational clusters were more likely to be in more deprived areas and were more likely to occur in areas of high population density. The distribution of multi-generational clusters varied according to ethnicity, with a higher proportion of multi-generational clusters that occurred in households with more than one ethnicity present (39.0%) than occurred in non-multi-generational clusters (15.8%). Limited to single ethnicity clusters, differences in distribution were noted by ethnicity. Solely-White households formed 72.4% of clusters in non-multi-generational clusters, compared to 44.5% in multi-generational households. Among multi-generational households, clusters in solely-Pakistani households formed 16.1% of clusters compared to 5.2% in non-multi-generational clusters, solely-Indian households formed 17.7% of clusters compared to 5.3% in non-multi-generational clusters and solely-Bangladeshi households 5.7% of clusters compared to 1.9% in non-multi-generational clusters.

### Trends in multi-generational clusters over time

The proportion of cases occurring in private residential dwellings has increased over the course of the pandemic (Table 2). Between 20 April 2020 and 14 June 2020, cases in private residential dwellings formed 72.7% of all cases, and by the period of 25 January 2021, cases in private residential dwellings formed 91.2% of all cases. Similarly, the percentage of cases occurring in private residential dwellings, that formed clusters increased over time. Between 20 April 2020 and 14 June 2020, clustered cases in residential properties formed 25.8% of all cases occurring in private residential dwellings, and by the period of 25 January 2021, clustered cases in private residential dwellings formed 50.7% of all cases occurring in private residential dwellings.

**Table 2:** Trend in cluster type over the course of the pandemic, England April 2020 to May 2021

| Epidemic period |  | Total cases |  |  | Non-multi-generational clusters |  | Multi-generational clusters |  | Per cent residential cases forming clusters (%) |
| --- | --- | --- | --- | --- | --- | --- | --- | --- | --- |
| Eight week period | Time span | All cases | Cases in residential properties | Per cent cases that were residential (%) | Clusters (n) | Cases (n) | Clusters (n) | Cases (n) |  |
| 0 | 20-04-2020 to 14-06-2020 | 118,361 | 86,095 | 72.7 | 9,644 | 21,942 | 68 | 301 | 25.8 |
| 1 | 15-06-2020 to 09-08-2020 | 37,647 | 28,502 | 75.7 | 3,953 | 10,013 | 79 | 375 | 36.4 |
| 2 | 10-08-2020 to 04-10-2020 | 193,057 | 152,207 | 78.8 | 25,122 | 64,783 | 280 | 1,311 | 43.4 |
| 3 | 05-10-2020 to 29-11-2020 | 951,953 | 827,556 | 86.9 | 164,428 | 403,620 | 2,036 | 11,846 | 50.2 |
| 4 | 30-11-2020 to 24-01-2021 | 1,811,348 | 1,667,937 | 92.1 | 384,326 | 1,002,442 | 6,834 | 42,783 | 62.7 |
| 5 | 25-01-2021 to 21-03-2021 | 530,797 | 484,167 | 91.2 | 96,235 | 240,629 | 1,104 | 4,927 | 50.7 |

Multi-generational clusters changed in prevalence over the course of the pandemic. In week 1 of 2020, multi-generational clusters formed only 1.39% of residential clusters (Figure 1). This increased over time, reaching a peak of 2.74% of residential clusters in week 32, but declined from that point on.

**Figure 1:**
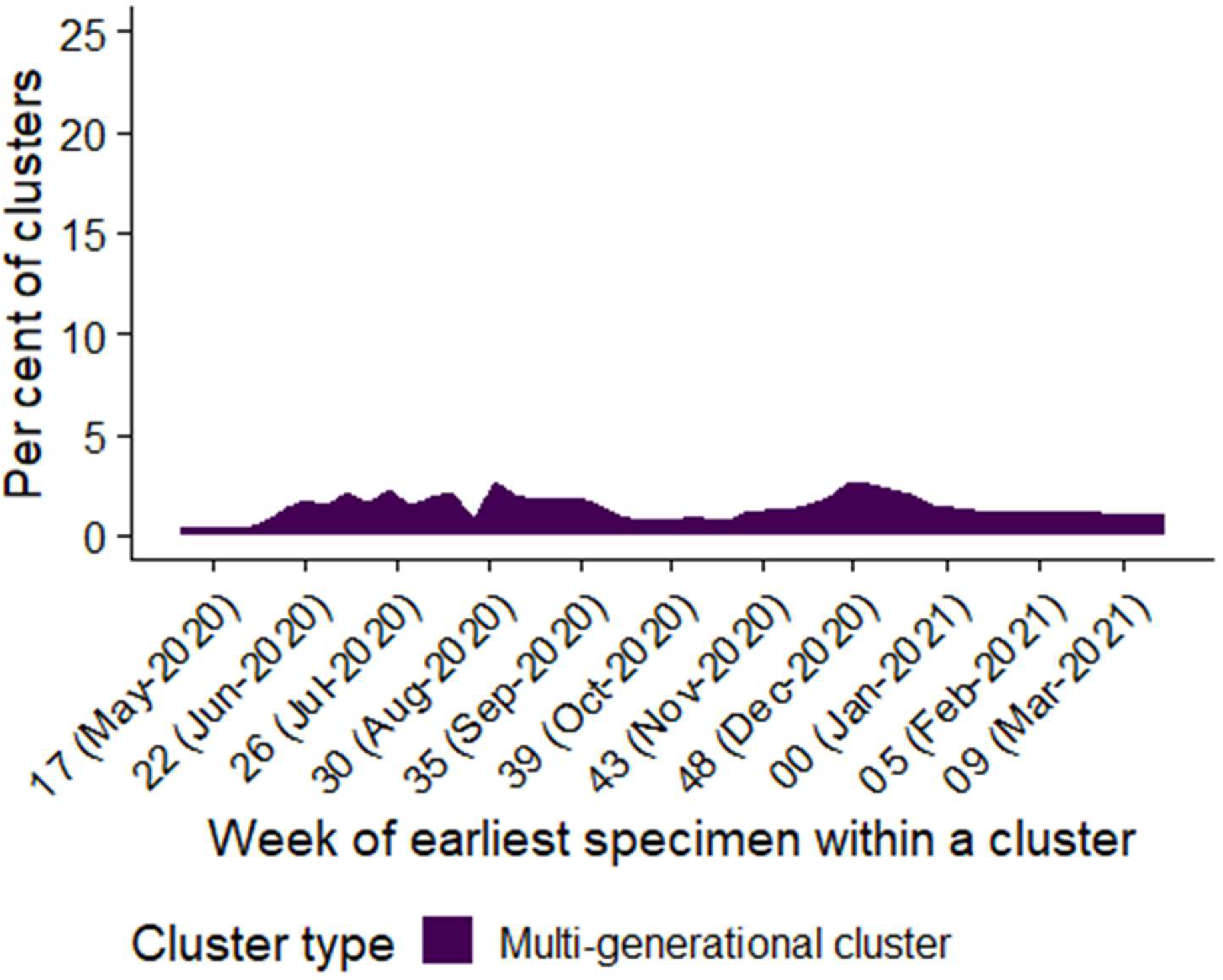
Weekly trend in cluster type occurring in residential dwellings, England

### Distribution of ethnicity among households with more than one ethnicity

As shown in table 1, 39.0% of multi-generational clusters occurred in households of with more than one ethnicity, compared to 15.8% of non-multi-generational clusters. To investigate the distribution of ethnicities further, the ethnicities of the cases in those clusters are shown in table 3.

**Table 3:** Distribution of cases in residential clusters by ethnicity in clusters occurring in mixed ethnicity, with at least three people in the household, England April 2020 to May 2021

| Ethnicity | Non-multigenerational clusters |  | Multigenerational clusters |  |
| --- | --- | --- | --- | --- |
|  | n cases | % | n cases | % |
| Asian Bangladeshi | 1,333 | 2.6 | 262 | 6.5 |
| Asian Indian | 2,881 | 5.6 | 286 | 7.1 |
| Asian other | 4,602 | 9 | 405 | 10 |
| Asian Pakistani | 2,503 | 4.9 | 265 | 6.5 |
| Black African | 2,619 | 5.1 | 301 | 7.4 |
| Black Caribbean | 1,527 | 3 | 150 | 3.7 |
| Black other | 982 | 1.9 | 83 | 2.0 |
| Mixed ethnicity | 5,105 | 10 | 309 | 7.6 |
| White British | 15,257 | 29.9 | 950 | 23.4 |
| White other | 9,381 | 18.4 | 618 | 15.3 |
| Other | 3,500 | 6.9 | 305 | 7.5 |
| Unknown | 1,345 | 2.6 | 118 | 2.9 |

Chi2 test for independence $p < 0.001$

People identified as White British were less commonly observed in mixed-ethnicity, multi-generational clusters (23.4%) than in non-multi-generational clusters (29.9%). In contrast, people identifying as Indian, Pakistani or Bangladeshi were more likely to be cases in multi-generational clusters than non-multi-generational clusters (7.1% vs 5.6%, 6.5% vs 4.9% and 6.5% vs 2.6% respectively, p < 0.001).

### Age of first case

To investigate how cases accrue in multi-generational clusters, we analysed the age of the first case within the cluster. Within multi-generational clusters, first cases were frequently 11-18 years of age (12.4% of multi-generational clusters, Table 4), 31-40 years of age (19.1%) or 61-70 years of age (15.3%).

**Table 4:** Distribution of cases in residential clusters by age group in multi-generational clusters and non-multi-generational clusters, England April 2020 to May 2021

| Age group | Non-multigenerational clusters |  |  | Multigenerational clusters |  |  |
| --- | --- | --- | --- | --- | --- | --- |
|  | Count first case | Count of subsequent cases | Per cent of cases that were first-cases | Count first case | Count of subsequent cases | Per cent of cases that were first-cases |
| 0-4 | 24,288 | 27,433 | 2.5 | 823 | 2,613 | 4.9 |
| 5-10 | 34,161 | 45,820 | 3.5 | 1,106 | 3,655 | 6.5 |
| 11-18 | 74,453 | 91,662 | 7.7 | 2,101 | 6,196 | 12.4 |
| 19-21 | 39,037 | 41,353 | 4.0 | 328 | 1,341 | 1.9 |
| 22-30 | 150,269 | 123,932 | 15.5 | 1,407 | 5,136 | 8.3 |
| 31-40 | 188,548 | 124,475 | 19.4 | 3,224 | 7,418 | 19.1 |
| 41-50 | 175,409 | 120,735 | 18.1 | 2,425 | 5,572 | 14.4 |
| 51-60 | 160,412 | 115,800 | 16.5 | 1,647 | 4,003 | 9.7 |
| 61-70 | 76,222 | 48,363 | 7.9 | 2,590 | 5,283 | 15.3 |
| 71-80 | 33,262 | 20,973 | 3.4 | 931 | 2,340 | 5.5 |
| ≥ 81 | 14,046 | 12,164 | 1.4 | 313 | 1,077 | 1.9 |
| Missing | 368 | 244 | <1 | 3 | 11 | <1 |

In comparison, among non-multi-generational clusters, first cases were distributed more around the middle of the range of ages, with 54.0% of first cases among those 31-60 years old. The distribution of first cases between the two settings reflects the general distribution for age for those settings (Figure 2).

**Figure 2:**
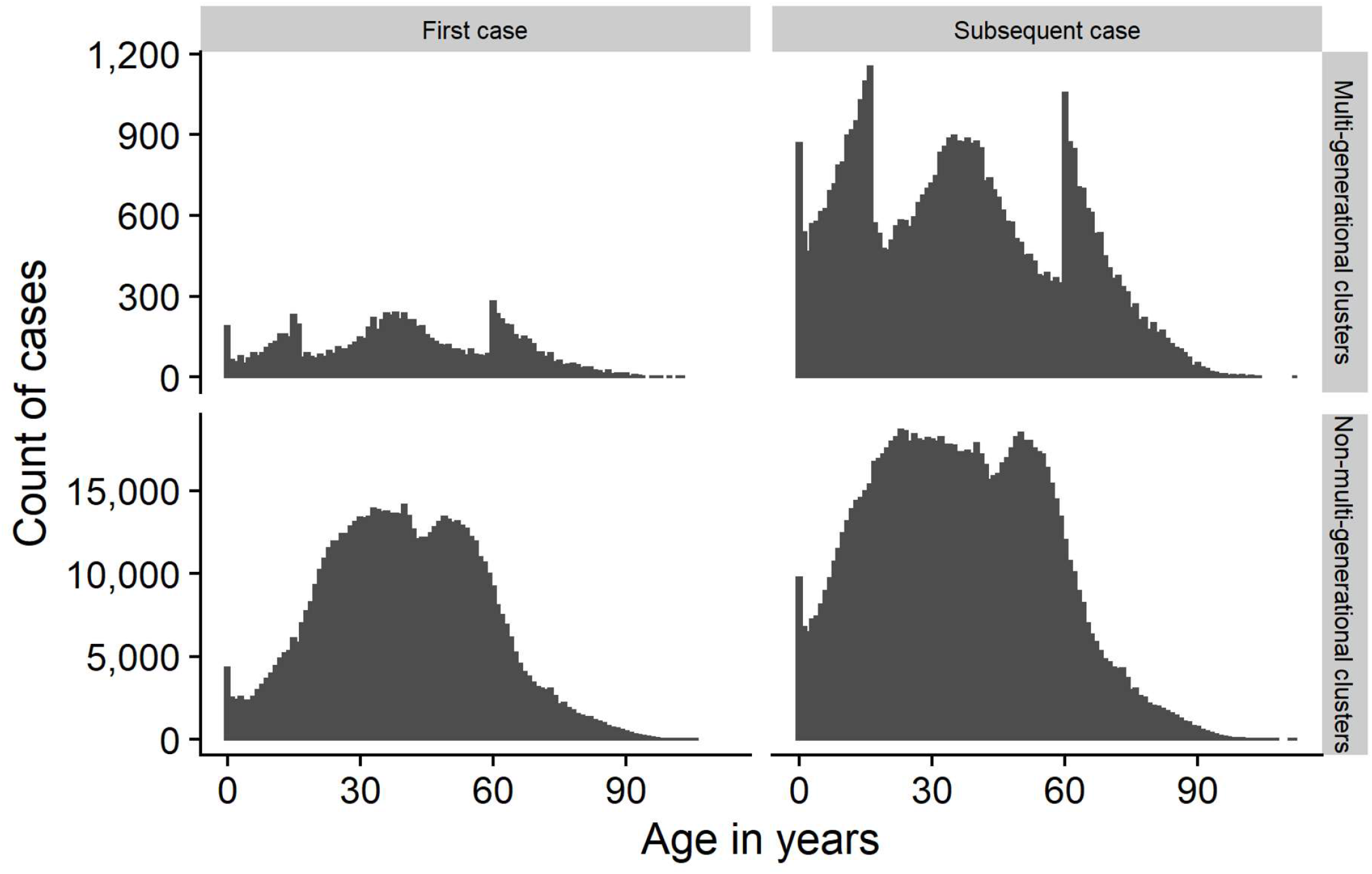
Distributions of cases occurring in residential dwelling clusters, by multi-generational status and first-case status. England April 2020 to May 2021

## DISCUSSION

We found that a minority of COVID-19 clusters occurred in multi-generational households, and at most only accounted for 2.7% of residential clusters (Figure 1). Therefore, the evidence gathered here do not support the hypothesis set out at the initiation of this study, namely that multi-generational households formed a large component of cases in the summer months in England (Table 2). Prior to these findings, a review from the UK government’s Scientific Advisory Group for Emergencies (SAGE) found that large or multi-generational households were associated with increased risk of transmission of COVID-19.[2] This acknowledges that there is an increased risk of transmission of COVID-19 within multi-generational households, although overall they may constitute a smaller proportion of the national cases and therefore rates of COVID-19 if the proportion of the population living in multi-generational households is low nationally. Supporting this argument is research published in May 2020 found that in 2013/14, only 1.7% of households in England had a grandparent residing in the same household.[4]

A major challenge is the lack of a consistent definition for multi-generational households, with different agencies collecting data and researching this area according to different definitions. Therefore, a working definition was used in this analysis to identify households with broad generational mix beyond more common parent-child family groupings.

Unfortunately, we only had access to data for people who tested positive for SARS-CoV-2, and not the number of household members. Therefore, we were unable to estimate secondary attack rates.

Despite this, several interesting inferences can be drawn from these data. Clusters of COVID-19 occurring in multi-generational households were more likely to be located in local areas of greater population density and relative deprivation than clusters of COVID-19 occurring in non-multi-generational households (Table 1). In addition, clusters of COVID-19 occurring in multi-generational households were less likely to be of solely-White ethnicity.

Therefore, the data are consistent with other findings which have shown than greater local-area deprivation is associated with higher rates of COVID-19 and that rates of diagnosis were higher among non-White ethnicities. [10] Similarly these data are consistent with findings that higher levels of population density are associated with higher rates of COVID-19 infection.[11]

Data from the English Housing Survey found that overcrowding is more common among non-White ethnicities, even after taking into account of income, suggesting that some of the drivers of overcrowding are cultural, and not simply economic.[12] The same data showed that Bangladeshi, Pakistani and Black African households were more likely to be overcrowded. This is not fully consistent with the data presented here, where multi-generational clusters were more common among Indian, Pakistani and Bangladeshi households.

### Age of first case

We found that the index case in multi-generational households clusters was infrequently a child less than 10 years. This finding is consistent with other, similar work; OpenSAFELY found living with children age 0-11 years old to not be associated with increased risk of infection, while other work found that healthcare workers with children less than 11 years old had a lower hazard rate of COVID-19 than healthcare workers without young children. [13,14] Also, a study of complete households across England found that households with children did not have different rates of transmission of COVID-19 than adult-pair households and demonstrated that index cases below the age of 15 years did not cause secondary cases in a high proportion of contacts. [15]

Together, this suggests that the risk of transmission from children to other members of the household is low, in turn suggesting that risk of transmission within multi-generational households is likely to occur between adults.

Although the number of cases in multi-generational clusters represents smaller absolute numbers in the pandemic, by definition, multi-generational households have high numbers of potential contacts, increasing opportunities for transmission. Government guidelines do not currently provide specific advice regarding managing COVID-19 in multi-generational households but advises that to limit close contact with others in a household if one has COVID-19 as little time as possible is spent in shared areas; that separate bathrooms are used; and that strict social distancing is observed. [16]

## Data Availability

Due to the highly personal nature of the data used in this study, data are not available.

## ACKNOWLEDGEMENTS

We thank the UKHSA GIS team for support in address matching cases.

## ETHICAL APPROVAL STATEMENT

As a product from routine public health surveillance of COVID-19, approval for the work was not sought from an Ethics Committee.

